# Incretin mimetics for weight loss forgive nonadherence

**DOI:** 10.1101/2025.03.22.25324451

**Authors:** Anıl Cengiz, Calvin C. Wu, Sean D. Lawley

## Abstract

**Aims:** GLP-1 and GIP-GLP-1 agonists have emerged as potent weight loss medications. These incretin mimetics often have low patient adherence, and as with any medication, clinically meaningful efficacy requires adequate adherence. But what constitutes “adequate” adherence for incretin mimetics? The purpose of this paper is to address this question.

**Materials and Methods:** We use mathematical modeling and stochastic simulation to investigate the weight loss efficacy of incretin mimetics under imperfect adherence. We use validated pharmacokinetic and pharmacodynamic models of semaglutide and tirzepatide and assume that simulated patients randomly miss doses.

**Results:** We find that semaglutide and tirzepatide forgive nonadherence, meaning that strong weight loss efficacy persists despite missed doses. For example, taking 80% of the prescribed doses yields around 90% of the weight loss achieved under perfect adherence. Taking only 50% of prescribed doses yields nearly 70% of the weight loss of perfect adherence. Furthermore, such nonadherence causes only small fluctuations in body weight, assuming that patient do not typically miss more than several consecutive doses.

**Conclusion:** Incretin mimetics are powerful tools for combating obesity, perhaps even if patients can consistently take only half of their prescribed doses. The common assumption that significant weight loss requires at least 80% adherence needs revision.

## 1 Introduction

Obesity is arguably the most urgent public health challenge today. Defined as having a Body Mass Index (BMI) in kg*/*m^2^ greater than or equal to 30, obesity is the most common chronic disease worldwide [1], and roughly 41% of adults in the United States (US) are obese [2]. Obesity increases mortality and the risk for other chronic diseases, such as type 2 diabetes, cardiovascular diseases, and certain cancers [3–5]. From an economic standpoint, obesity costs over $170 billion in healthcare spending in the US every year [6].

Incretin mimetics, such as glucagon-like peptide-1 (GLP-1) and dual GLP- 1/gastric inhibitory polypeptide (GLP-1/GIP) receptor agonists, have emerged as potent treatments for obesity [7, 8]. Indeed, semaglutide and tirzepatide respectively induce placebo-adjusted average body weight losses of 15% and 19% [9], which far exceeds the 3% to 7% weight loss typically associated with prior generations of weight loss drugs [10].

Adequate medication adherence is necessary to achieve substantial weight loss from incretin mimetics. Medication adherence, which is the process by which patients take their medications as prescribed [11], can be divided into the three phases of initiation (starting treatment), implementation (actual dosing follows the prescribed regimen), and persistence (continuing treatment) [12]. Poor adherence has been reported for incretin mimetics [13–15], with claims data suggesting that perhaps only around half of patients take at least 80% of their doses while on treatment and less than one third of patients continue treatment past one year [16]. High prices and supply shortages of incretin mimetics may have contributed to these poor adherence rates [17]. More generally, however, nonadherence is a well-documented problem for many classes of medications [18, 19]. Indeed, nonadherence is especially problematic for long-term pharmacotherapies for chronic conditions [20].

The implementation phase of adherence is often measured from electronic healthcare databases in terms of the proportion *p* of days covered, which approximates the proportion of prescribed doses that the patient actually takes [21]. Originally estimated for antihypertensive medications [22], the threshold *p* = 80% has been widely adopted to distinguish adherent (*p*≥ 80%) versus nonadherent (*p <* 80%) patients across many classes of medication [23]. Indeed, for incretin mimetics, Mody et al. [13] and Gleason et al. [16] both used the *p* = 80% threshold to define adherent patients. Furthermore, the recent statistical modeling work of Pandey et al. [2] assumed that significant weight loss occurs only for patients with adherence greater than *p* = 80%.

How does weight loss from incretin mimetics depend on adherence? What is an “adequate” adherence? If a patient takes only *p* = 80% of their prescribed doses, how much weight will they lose? What if they only take *p* = 50% of their prescribed doses? How does this depend on the specific incretin mimetic and dose size?

The purpose of this paper is to address these questions using mathematical modeling and stochastic simulation. We consider semaglutide (brand name Wegovy for weight loss and Ozempic and Rybelsus for type 2 diabetes) and tirzepatide (brand name Zepbound for weight loss and Mounjaro for type 2 diabetes). We use recent pharmacokinetic (PK) and pharmacodynamic (PD) models of semaglutide [24] and tirzepatide [25], which accurately reproduce the weight loss observed in clinical trials [7, 8]. We model patient adherence by a stochastic process and analyze the resulting weight loss. In particular, we assume that patients miss doses randomly and compare the induced weight loss to perfectly adherent patients. The concept is illustrated in Figure 1, which shows sample PK time courses (drug concentration in left panel) and PD time courses (weight loss in right panel) for perfect adherence (red) and imperfect adherence (blue). Our results are obtained by averaging many such stochastic samples.

**Figure 1:**
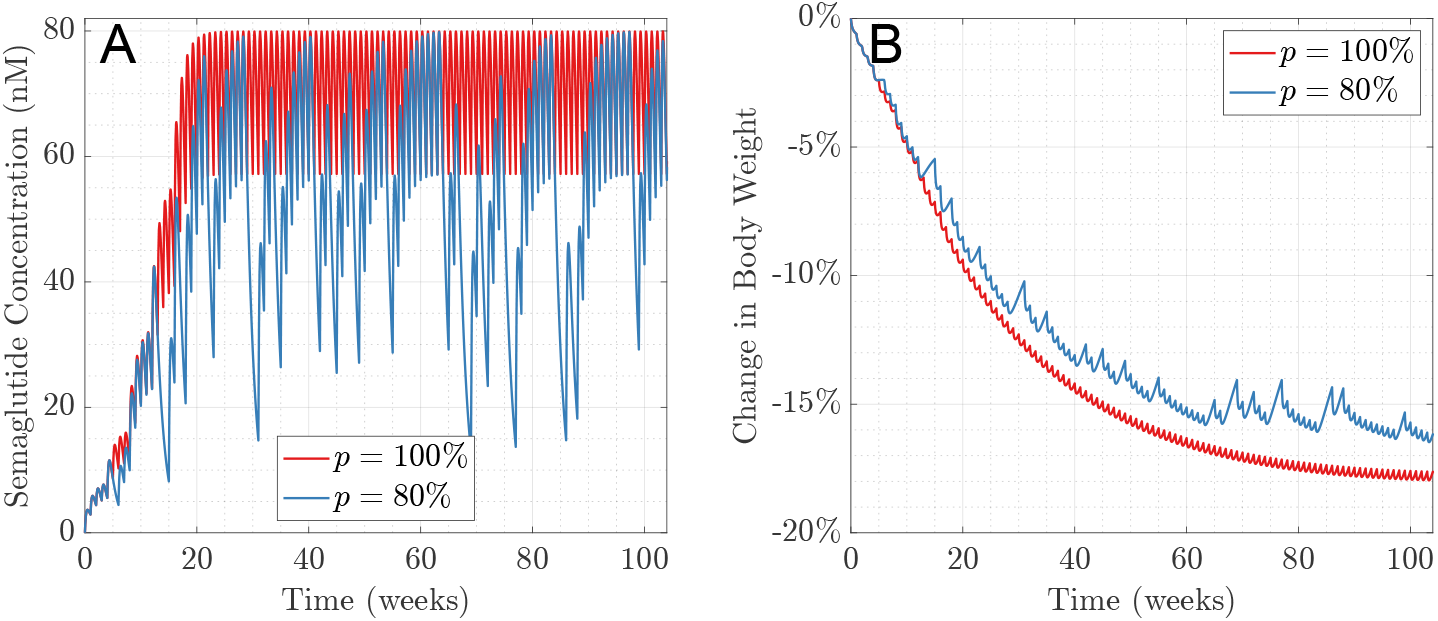
Sample time courses for semaglutide concentration (left panel) and change in body weight (right panel). The red curves correspond to perfect adherence with *p* = 100%, and the blue curves correspond to *p* = 80% adherence.

Our analysis predicts that semaglutide and tirzepatide are highly forgiving [18], meaning that they maintain high weight loss efficacy despite lapses in patient adherence. For example, we find that patients who take *p* = 80% of their doses retain roughly 90% of the weight loss achieved under perfect adherence (i.e. *p* = 100%). Furthermore, taking *p* = 50% of doses yields nearly 70% of the weight loss achieved under perfect adherence. We also determine clinical scenarios in which poor adherence to a higher dose yields greater weight loss than perfect adherence to a lower dose. For example, we find that *p >* 60% adherence to a 10 mg tirzepatide dosing regimen yields more weight loss than *p* = 100% adherence to a 5 mg tirzepatide dosing regimen.

## 2 Methods

### 2.1 PKPD models

We use PKPD models to describe the effects of semaglutide and tirzepatide. The full PKPD details (equations and parameter values) are given in [25].

For semaglutide, we use the PKPD model of Strathe et al. [24]. The PK model is a standard one compartment model with first-order absorption and elimination. PD is a semi-mechanistic, non-linear exposure-response model. This PKPD model recapitulates longitudinal weight loss data from three randomized, double-blind, controlled trials of subcutaneous semaglutide [8, 26, 27]. For tirzepatide, we use the PKPD model of [25]. The PK model is a standard two compartment model with first-order kinetics, which was first proposed by the US Food and Drug Administration [28]. The PD model has the same structure as the semaglutide PD model of Strathe et al. [24], but was reparameterized to fit the longitudinal weight loss data from the double-blind, randomized, controlled trial of Jastreboff et al. [7].

### 2.2 Adherence models

We assume that a simulated patient is prescribed a once-weekly dose of either 2.4 mg of semaglutide or 5, 10, or 15 mg of tirzepatide. We model the patient’s adherence to this prescribed regimen by a stochastic process.

Specifically, we assume that the patient takes a given weekly dose with probability *p* and otherwise misses the weekly dose. Furthermore, we let *q* denote the correlation coefficient between successive prescribed doses. That is, if *q* = 0, then missing doses are independent, i.e. missing one dose does not make the patient any more or less likely to miss the next dose. Further, if *q >* 0, then missing one dose makes the patient more likely to miss the next dose. Finally, if *q <* 0, then missing one dose makes the patient less likely to miss the next dose. In effect, the value of *p* determines how many doses are taken or missed over a long period of time, and the value of *q* determines if missed doses tend to occur either consecutively (*q >* 0), separated by taken doses (*q <* 0), or uniformly (*q* = 0).

To make our model of patient adherence precise, we need more mathematical notation. Following [29], the patient’s dose taking is described by a sequence of identically distributed Bernoulli random variables {*ξ*_*n*_}_*n*_, where

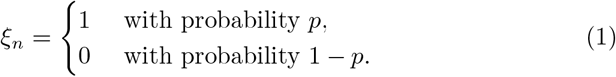

In particular, *ξ*_*n*_ = 1 means that the patient took dose *n*, and *ξ*_*n*_ = 0 means that the patient missed dose *n*. We assume that {*ξ*_*n*_}_*n*_ is a two-state Markov chain with the following transition probabilities [30],

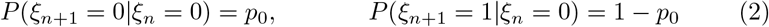

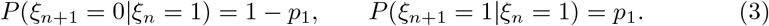

In words, (2) means that if the patient misses a dose, then they have probability *p*_0_ of missing the next dose. Similarly, (3) means that if the patient takes a dose, then they have probability *p*_1_ of taking the next dose. The long-term fraction of prescribed doses that are actually taken by the patient is [29]

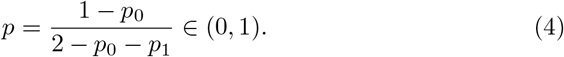

The correlation coefficient between any two successive doses is [29]

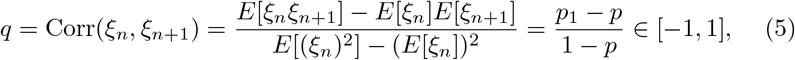

where *E* denotes mathematical expectation. In general, correlation coefficients range from *q* = −1 to *q* = 1, but fixing the value of *p* ∈ (0, 1) restricts the range of *q*. In particular,

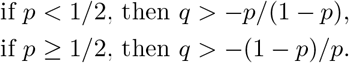

Note that while (4)-(5) give *p* and *q* in terms of *p*_0_ and *p*_1_, we can equivalently write *p*_0_ and *p*_1_ in terms of *p* and *q*,

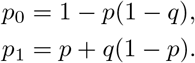

To gain intuition for the meaning of a particular choice of *q*, it is useful to consider the average number of consecutive missed doses, which we denote by *t*_0_. Given that one dose is missed, the expected number of additional missed doses is a geometric random variable *G* ∈ {0, 1, 2, …} with mean *p*_0_*/*(1 − *p*_0_). Therefore, the expected number of consecutive missed doses is *t*_0_ = 1*/*(1− *p*_0_), and therefore (4)-(5) imply that

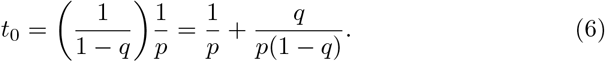

Notice that

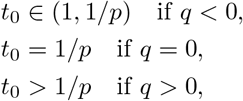

and lim_*q*→1_ *t*_0_ = ∞. Another useful quantity is the average number of consecutive taken doses, which we denote by *t*_1_. By the analogous argument to obtain *t*_0_, we have that *t*_1_ = 1*/*(1 − *p*_1_), and therefore (4)-(5) imply that

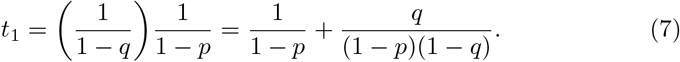

Note that this adherence model can equivalently parameterized in terms of either the pair (*p*_0_, *p*_1_), or the pair (*p, q*), or the pair (*t*_0_, *t*_1_).

For example, suppose *p* = 80%. If *q* = 0, then *t*_0_ = 1.25 and *t*_1_ = 5, and thus patients typically take around 5 consecutive doses before an isolated missed dose. If *q* = 0.6, then *t*_0_ = 3.125 and *t*_1_ = 12.5, and thus patients tend to take around 12 or 13 consecutive doses before missing around three doses in a row. Importantly, however, patients still take *p* = 80% of their doses over time regardless of *q*.

### 2.3 Weight loss metrics

Define the relative body weight reduction,

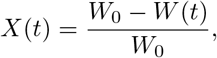

where *W*_0_ denotes the patient’s body weight at the start of treatment and *W* (*t*) denotes the patient’s body weight after being on the treatment for time *t* ≥ 0. We use the superscript “perf” on *X* to indicate a perfectly adherent patient, i.e. *X*^perf^, which corresponds to setting *p* = 1. In addition, we use subscripts on *X* to indicate the drug and dose size, i.e. *X*_semag_, *X*_tirz,5mg_, *X*_tirz,10mg_, and *X*_tirz,15mg_, denote the relative body weight reduction for a patient prescribed semaglutide at 2.4 mg per week, and tirzepatide at 5 mg, 10 mg, and 15 mg per week, respectively.

For the relative body weight reduction time course {*X*(*t*))_*t*≥0_, let ⟨*X*⟩ denote the long-term average,

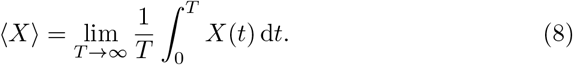

The standard deviation of *X* is denoted by SD(*X*) and defined by

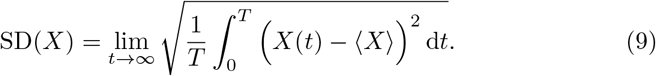

The fraction of time that *X* is above a level *x* is

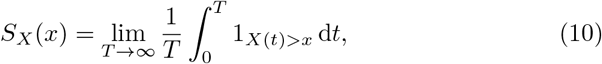

where 1_*X*(*t*)*>x*_ denotes the indicator function on the event *X*(*t*) ≤ *x* (i.e. 1_*X*(*t*)*>x*_ = 1 if *X*(*t*) *> x* and 1_*X*(*t*)*>x*_ = 0 if *X*(*t*) ≤ *x*). In numerical simulations, we approximate the limits in (8)-(10) by taking *T* = 200 years (for computational efficiency, we take *T* = 100 years for simulations with nonzero correlations in missed doses).

## 3 Results

We now use the models and metrics of section 2 to investigate the effects of nonadherence to semaglutide and tirzepatide. In sections 3.1–3.3, we assume that patients miss each dose randomly and independently of their prior doses, i.e. missing a dose one week does not make the patient any more or less likely to miss a dose the following week. We relax this independence assumption in section 3.4 and consider the effects of positive or negative correlations in missed doses.

### 3.1 Average weight loss

Figure 2A plots the average weight loss (denoted ⟨*X*⟩ and defined in (8)) as a function of the proportion of doses taken (denoted *p*) over a long period of time. Figure 2 predicts that semaglutide and tirzepatide are very “forgiving” [31] in that substantial weight loss is achieved despite lapses in missed doses. For instance, Figure 2A predicts that a patient prescribed a once-weekly dose of either 2.4 mg of semaglutide or 5 mg of tirzepatide maintains around a 15% reduction in body weight if they tend to miss about one dose out of every 5 doses, i.e. *p* = 80%. Furthermore, at this same *p* = 80% level of adherence, a once-weekly prescription of either 10 or 15 mg of tirzepatide yields 20% or 23% weight loss, respectively.

**Figure 2:**
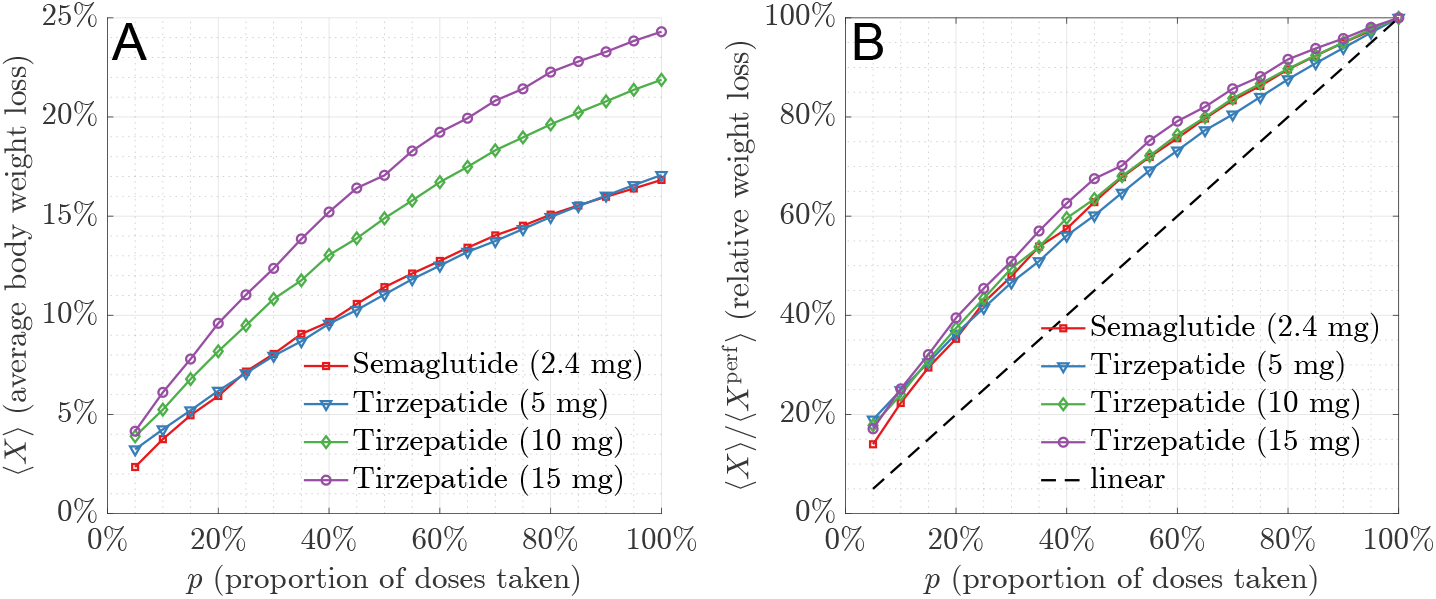
Missed doses do not proportionately decrease efficacy. Panel A plots the average body weight loss (denoted ⟨*X*⟩ and defined in (8)) as a function of adherence *p* for different drugs and dose sizes. Panel B shows how missed doses reduce weight loss relative to perfect adherence.

Furthermore, Figure 2A shows that lower adherence to a higher dose may yield greater weight loss than perfect adherence to a lower dose. For instance, perfect adherence (i.e. *p* = 100%) to either 2.4 mg of semaglutide or 5 mg of tirzepatide produces about 17% weight loss. However, this same 17% weight loss is achieved with an adherence of only *p* = 60% to a 10 mg regimen of tirzepatide or adherence *p* = 50% to a 15 mg regimen of tirzepatide.

Figure 2B compares the weight loss achieved with imperfect adherence versus perfect adherence. Naively, one might expect that taking, say, *p* = 75% of doses would yield 75% of the weight loss achieved under perfect adherence. The black dashed line in Figure 2B illustrates this simple linear expectation. The main point of Figure 2B is that the relative weight loss for semaglutide and tirzepatide is considerably above this linear expectation. For instance, taking *p* = 75% of prescribed doses yields around 80 to 85% of the weight loss achieved under perfect adherence. Furthermore, taking *p* = 50% of prescribed doses yields around 65 to 70% of the weight loss achieved under perfect adherence.

The strong “forgiveness” shown in Figure 2 is not surprising in light of the clinical data on both semaglutide [26] and tirzepatide [7]. Indeed, the standard commercial dosing regimens of these incretin mimetics appear to be at the top end of a saturating dose-response curve, which means that increasing or decreasing the dose does not proportionately affect weight loss. For instance, tripling the weekly tirzepatide dose from 5 mg to 15 mg constitutes a 200% dose increase but increases weight loss by less than 50% [7]. Based on this clinical data, one might predict that taking only one third of prescribed doses (i.e. *p* = 33%) would decrease actual weight loss by a mere 50%. That is, one might expect that the relative weight loss is

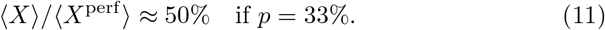

Figure 2B shows that the simple prediction in (11) is quite accurate, though (11) slightly underestimates the relative weight loss achieved for higher dose regimens such as 15 mg of tirzepatide.

### 3.2 Body weight fluctuations

Section 3.1 considers only the long-term average weight loss, and thus ignores how missed doses cause fluctuations in body weight. To illustrate such fluctuations, Figure 3A plots the change in body weight time course for a perfectly adherent patient (*p* = 100%, red) and an imperfectly adherence patient (*p* = 80%, blue) over several years of semaglutide therapy (this plot omits the beginning of treatment, which is illustrated in Figure 1). Notice that the perfectly adherent patient’s body weight only negligibly fluctuates. While the imperfectly adherent patient experiences much greater body weight fluctuations, their body weight still only rarely deviates more than about 1% or 2% from their average of ⟨*X*_semag_⟩ = 15%.

**Figure 3:**
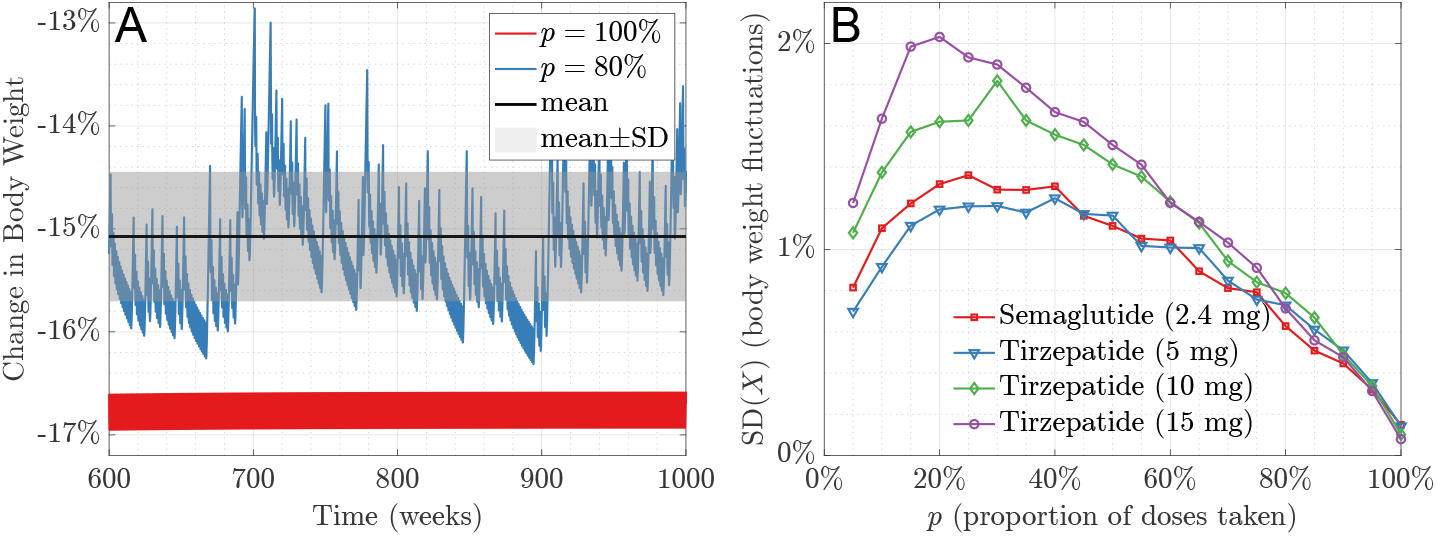
Body weight fluctuations. Panel A shows that the body weight of an imperfectly adherent patient rarely deviates by more than a few percentage points from their long-term average. Panel B quantifies such fluctuations by plotting the standard deviation of relative body weight loss (denoted SD(*X*) and defined in (9)) as a function of adherence *p* for different drugs and dose sizes.

The standard deviation of relative body weight reduction, denoted SD(*X*) and defined in (9), quantifies such fluctuations. Roughly speaking, SD(*X*) is the average deviation from the mean and is illustrated by the gray region in Figure 3A. Notice that the body weight reduction is typically confined to the region spanned by the mean plus or minus one standard deviation (gray region), and only very rarely fluctuates more than two or three standard deviations from the mean.

While Figure 3A shows an example weight loss time course, Figure 3B plots SD(*X*) as a function of the adherence *p* for different drugs and dose sizes. Notice that SD(*X*) is never greater than 2% and is closer to 1% for most adherence levels *p* and dose sizes. Such minor fluctuations may be clinically insignificant, and indeed they are similar to typically daily, weekly, and seasonal fluctuations in the absence of incretin mimetics [32].

### 3.3 More detailed weight loss statistics

A more detailed statistic of body weight reduction is the fraction of time that a patient experiences weight loss greater than a given level. For instance, we have seen that a patient with *p* = 80% adherence to semaglutide tends to have their body weight reduction between 14 and 16% most of the time. To make this more precise, we defined *S*_*X*_ (*x*) in (10), which is the fraction of time that weight loss is greater than a given level *x*. This quantity allows us to determine, for instance, how often a patient’s weight loss is greater than *x* = 10% of their baseline body weight.

Figure 4 plots *S*_*X*_ for different drugs, dose sizes, and adherence rates *p*. Notice that Figure 4A shows that a patient with *p* = 80% adherence to semaglutide essentially always has a weight loss greater than 80% of the weight loss they would have achieved with perfect adherence. In fact, examining Figure 4 shows that this pattern extends to all doses of tirzepatide and all adherence rates *p* considered. That is, a given adherence *p* ensures that a patient will nearly always have weight loss greater than *p* ⟨*X*^perf^⟩. For example, *p* = 40% adherence to 15 mg of tirzepatide will almost always have weight loss greater than 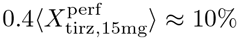 at all times (and we know from Figure 2A that the average weight loss for such a patient is ⟨*X*_tirz,15mg_⟩ ≈ 15%). Though greater degrees of weight loss are generally associated with greater health improvements for people with obesity, a wide range of cardiometabolic benefits are seen at 10% weight loss, and substantial health gains are present at even around 5% weight loss [33].

**Figure 4:**
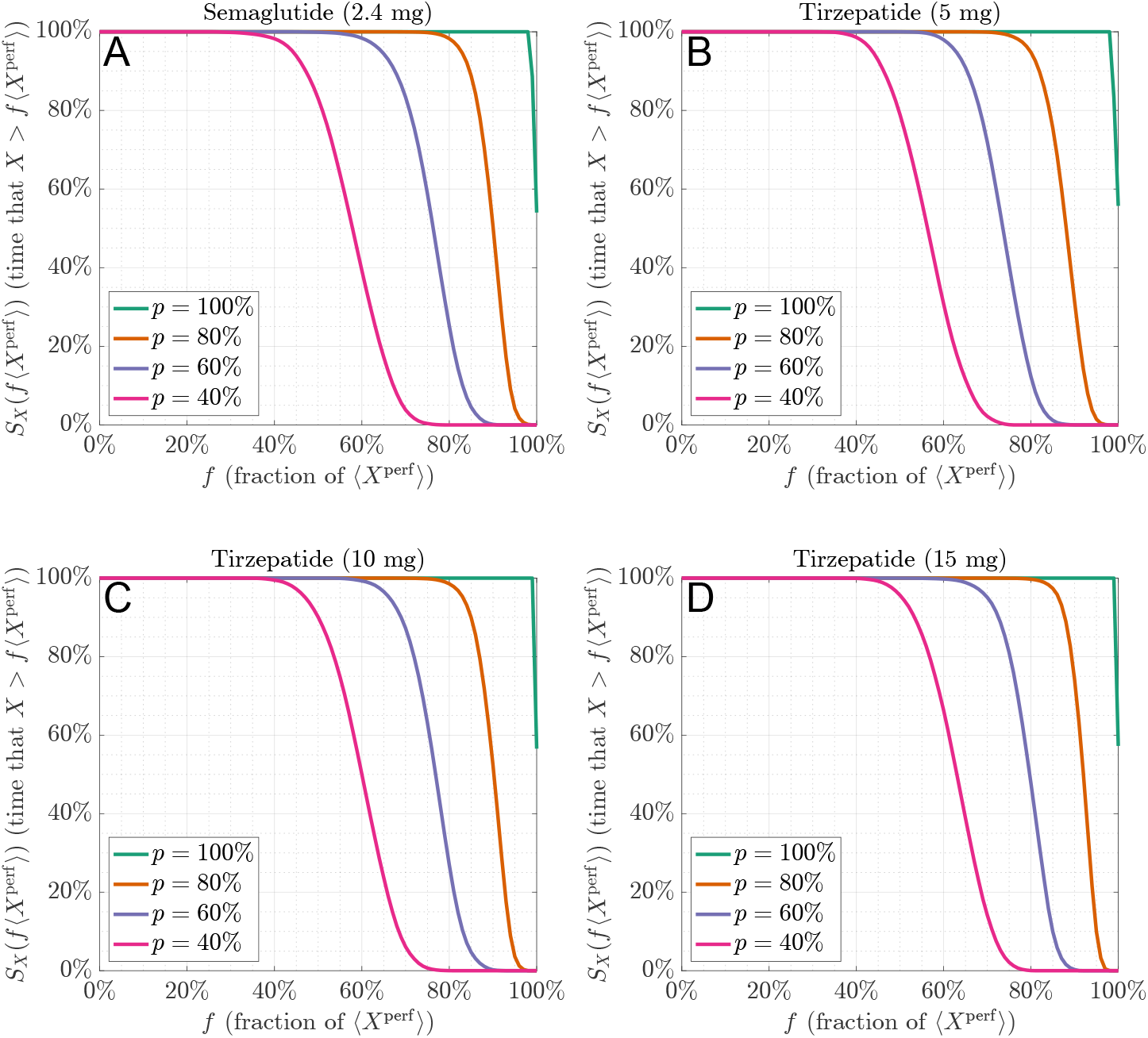
Plots of *S*_*X*_ in (10) for different drugs and dose sizes. In words, *S*_*X*_ (*f* ⟨*X*^perf^⟩) is the fraction of time that a patient will have weight loss greater than a fraction *f* of the weight loss that they would achieve under perfect adherence.

### 3.4 Correlations in missed doses

The results in sections 3.1–3.3 and Figures 2–4 assume that there are no correlations in missed doses (i.e. *q* = 0). That is, sections 3.1–3.3 assume that patients are no more or less likely to miss a dose following a missed dose. We now relax this assumption so that after a missed dose, the patient may be more likely (*q >* 0) or less likely (*q <* 0) to miss the next dose. Figure 5 shows how the average weight loss with imperfect adherence, ⟨*X*⟩, compares to the average weight loss with perfect adherence, ⟨*X*^perf^⟩ for *q >* 0 and *q <* 0 (analogous to Figure 2B). This figure shows that positive correlations in missed doses (which means that missed doses tend to occur consecutively) reduces average weight loss. However, Figure 5 shows that incretin mimetics are still quite forgiving of missed doses even if *q >* 0. For instance, if *p* = 80% and *q* = 0.6, then patient still maintain more than 85% of the weight loss of perfect adherence. Note that setting *q* = 0.6 for *p* = 80% describes the rather extreme case that patients tend to go an entire month with taking any medication (see (6)-(7)).

**Figure 5:**
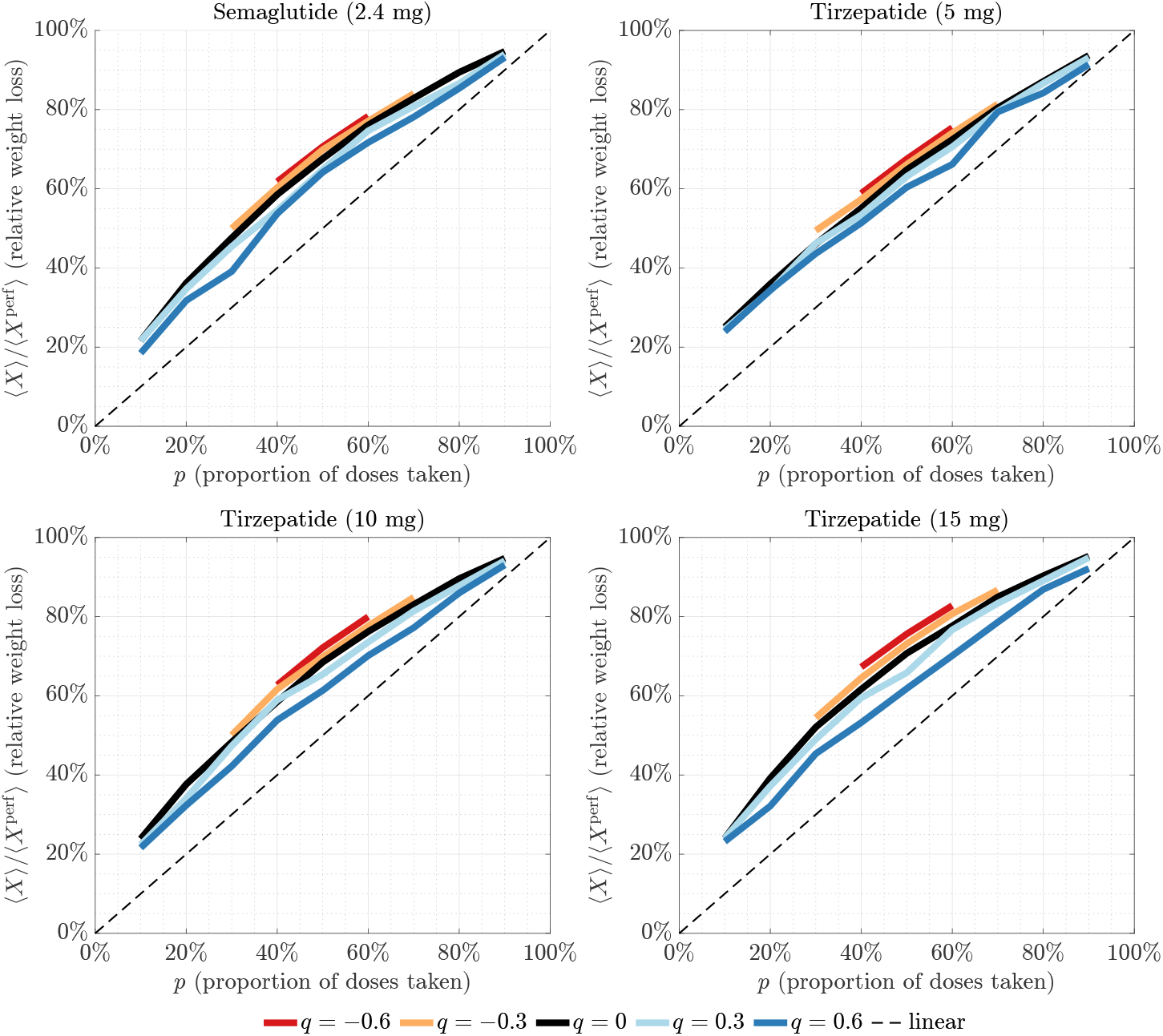
Weight loss relative to perfect adherence for positive (*q >* 0) or negative (*q <* 0) correlations in missed doses.

## 4 Discussion

In this paper, we used mathematical modeling to investigate the weight loss efficacy of incretin mimetics when patients sporadically miss doses. Our analysis predicts that semaglutide and tirzepatide forgive missed doses, meaning that they maintain strong weight loss efficacy under imperfect adherence. In particular, patients can maintain significant average weight loss over time with relatively small body weight fluctuations even when they take only around half of their doses.

We took existing PKPD models and investigated their response to imperfect patient adherence modeled by a stochastic process. This basic approach has been used to investigate a number of questions related to medication adherence, including taking a double dose after a missed dose [34], taking or skipping a late dose [35], and the effects of nonadherence to antihypertensives [29, 36], antiepileptics and antipsychotics [37], antidiabetics [38], and antibiotics [39]. Mathematical modeling has emerged as an important tool for studying non-adherence, as it avoids the ethical issues inherent to forcing real patients to miss doses [40] by leveraging the established science of pharmacometrics and the theory of stochastic processes.

Since our investigation was based on mathematical models, it ultimately awaits empirical validation. Nevertheless, our main result that semaglutide and tirzepatide forgive missed doses agrees with data from clinical trials [7, 8, 26]. To elaborate, the standard dosing regimens of these incretin mimetics appear to be at the top end of a saturating dose-response curve, meaning that increasing or decreasing the dose does not proportionately affect weight loss. This is evident from comparing the relatively small additional weight loss from increasing the weekly dose of (i) semaglutide by 140% (1 mg to 2.4 mg) [8, 26] or (ii) tirzepatide by 100% (5 mg to 10 mg) or 50% (10 mg to 15 mg) [7]. Based on these diminishing returns observed in the clinic, one should expect that missing some given percentage of doses does not proportionately decrease weight loss, which is exactly what we found. Our prediction that semaglutide and tirzepatide are forgiving also agrees with the clinical experience of the second author (detailed in the recent case report [41]), which was the original motivation for this study.

Naturally, our mathematical model made a number of simplifying assumptions. For one, we assumed that each dose was either taken on the prescribed day or missed entirely. That is, we did not allow for “late” doses that are taken, say, 10 days after the prior dose. While a more detailed model could incorporate such late doses, we expect that such an analysis would only show semaglutide and tirzepatide to be even more forgiving, since taking a dose later than scheduled is surely more efficacious than skipping the dose entirely. We also assumed a simple stochastic model of adherence parameterized by only the fraction of doses taken *p* and correlation between doses *q*. However, the human behavioral process of medication adherence is assuredly more complicated. Furthermore, while our allowance for correlations in missed doses via the parameter *q* can model such phenomena as “drug holidays” [42], we are not aware of publicly available data specific to incretin mimetics that could be used to precisely parameterize such correlations (i.e. to choose *q*). This underscores the need for more detailed patient adherence data, rather than merely the fraction of doses taken *p*. Nevertheless, we found that incretin mimetics remain forgiving even for very strong correlations in which patients typically go an entire month with taking any medication.

These limitations notwithstanding, the preponderance of evidence suggests that semaglutide and tirzepatide maintain strong weight loss efficacy in spite of missed doses. In particular, it appears that occasional missed doses are very unlikely to nullify treatment with incretin mimetics. The assumption that *p* = 80% is necessary for significant weight loss [2, 13, 16] is likely too pessimistic and needs revision. This contrasts “unforgiving” drugs, in which missing just 10% of doses can cause treatment failure [43]. This work is not intended to discourage patients and healthcare providers from striving for high adherence. Rather, the predicted forgiveness of incretin mimetics is heartening news that treatment should not be abandoned if patients cannot consistently take at least *p* = 80% of their prescribed doses. Indeed, our analysis predicts that this new generation of medications are powerful tools for combating obesity, perhaps even if patients take only half of their prescribed doses.

## Data Availability

All data produced in the present study are available upon reasonable request to the authors.

## Acknowledgments

SDL and AC were supported by the National Science Foundation (Grant Nos. CAREER DMS-1944574 and DMS-2325258).

## Notes

### Competing Interest Statement

The authors have declared no competing interest.

### Author Declarations

The study used ONLY openly available human data that were originally located at https://doi.org/10.1056/NEJMoa2206038 and https://doi.org/10.1056/NEJMoa2032183.

